# Increased household secondary attacks rates with Variant of Concern SARS-CoV-2 index cases

**DOI:** 10.1101/2021.03.31.21254502

**Authors:** Sarah A. Buchan, Semra Tibebu, Nick Daneman, Michael Whelan, Thuva Vanniyasingam, Michelle Murti, Kevin A. Brown

**Affiliations:** Health Protection, Public Health Ontario, 661 University Ave., Floor 17, Toronto, ON, M5G 1M1, Canada

## Abstract

**IMPORTANCE:** Higher secondary attack rates related to variant of concern (VOC) index cases have been reported, but have not been explored within households, which continue to be an important source of coronavirus disease 2019 (COVID-19) transmission

**OBJECTIVE:** To compare secondary attack rates in households with VOC versus non-VOC index cases.

**DESIGN:** A retrospective cohort study of household index cases reported from February 7 – 27, 2021. A propensity-score matched cohort was derived to calculate adjusted estimates.

**SETTING:** Ontario, Canada.

**PARTICIPANTS:** A population-based cohort of all private households with index cases. We excluded cases in congregate settings, as well as households with one individual or with >1 case with the same earliest symptom onset date.

**EXPOSURE:** VOC status, defined as either individuals confirmed as B.1.1.7 using whole genome sequencing or those that screened positive for the N501Y mutation using real-time PCR.

**MAIN OUTCOME AND MEASURE:** Household secondary attack rate, defined as the number of household secondary cases that occurred 1-14 days after the index case divided by the total number of household secondary contacts.

**RESULTS:** We included 1,259 index VOC and non-VOC cases in the propensity score-matched analysis. The secondary attack rate for VOC index cases in this matched cohort was 1.31 times higher than non-VOC index cases (RR=1.31, 95%CI 1.14-1.49), similar to the unadjusted estimate. In stratified analyses, the higher secondary attack rate for VOC compared to non-VOC index cases was accentuated for asymptomatic index cases (RR=1.91, 95% CI 0.96-3.80) and presymptomatic cases (RR=3.41, 95%CI 1.13-10.26)

**CONCLUSIONS AND RELEVANCE:** This study provides strong evidence of increased transmissibility in households due to VOCs and suggests that asymptomatic and pre-symptomatic transmission may be of particular importance for VOCs. Our study suggests that more aggressive public health measures will be needed to control VOCs and that ongoing research is needed to understand mechanisms of VOC transmissibility to curb their associated morbidity and mortality.

## Introduction

The prevalence of variants having the N501Y mutation have rapidly increased globally,^1^ including in Ontario, Canada, where this prevalence increased dramatically in February, 2021. These patterns of rapid strain replacement suggest increased transmissibility of variants with the N501Y mutation, which is present across variants of concern (VOC), including B.1.1.7, B.1.351, and P.1 lineages.^2^ However the exact degree of increased transmissibility, and specific settings when increased transmissibility occurs, remains unclear. ^1, 3,4,5^ Higher secondary attack rates related to VOC index cases have been reported, ^6^ but have not been explored within households, which continue to be an important source of coronavirus disease 2019 (COVID-19) transmission.^7^ The objective of this study was to compare secondary attack rates in households with VOC versus non-VOC index cases in Ontario, Canada.

## Methods

We identified individuals with laboratory-confirmed COVID-19 reported in the Public Health Case and Contact Management Solution (CCM), Ontario’s COVID-19 reporting system, and included households with index cases reported from February 7 to 27, 2021. VOC cases included either individuals confirmed as B.1.1.7 using whole genome sequencing or those that screened positive for the N501Y mutation using real-time PCR. B.1.1.7 accounted for 94% of confirmed VOC cases reported in Ontario until February 27, 2021.^8^ All PCR-positive specimens in Ontario with cycle threshold ≤35 underwent screening for the N501Y mutation during the study period using real-time PCR. Non-VOC cases were those that screened negative for the N501Y mutation.

We grouped cases living in the same household based on residential address.^9^ Index cases were defined as the first case in the household based on symptom onset date (or specimen collection date, if symptom onset date was not available) and secondary cases were included if they occurred 1-14 days after the index case. We excluded cases in congregate settings, as well as households with one individual or with >1 case with the same earliest symptom onset date. We used reported household size to calculate secondary attack rates by dividing the number of secondary cases by the total number of household secondary contacts (i.e., household size minus one). Cases with and without symptoms were classified based on symptom information and onset date reported in CCM.

Poisson regression was performed for the unadjusted and adjusted analyses, overall and by strata. The models were specified with the count of household secondary cases as the outcome, the logarithm of household size as the offset and VOC status as a binary exposure covariate. Clustering was accounted for with a random intercept for household to account for known overdispersion.

Our unadjusted risk ratios (RR) included the entire cohort of households with a VOC or non-VOC index case in Ontario. Adjusted RRs were based on a propensity score matched analysis. VOC index cases were 1:1 matched to non-VOC index cases based on gender and age group, and matched on the logit of the propensity score, using a caliper width of 0.2 times the standard deviation.^10^ The propensity score was based on a logistic regression model of VOC status as a function of five covariates: reported date, time between symptom onset and testing, association with a reported outbreak, as well as the neighbourhood proportion of visible minority residents (non-white and non-Indigenous population) and household crowding as determined using 2016 Canadian Census data.^11^ Regression estimates were reported using RRs and 95% confidence intervals (CI). Statistical analysis was performed in R version 4.0.4.^12^

This study was approved by Public Health Ontario’s Research Ethics Board.

## Results

We identified 5,617 index cases and 3,397 secondary cases across the study period. Amongst index cases, 1,318 were classified as VOC (151 B.1.1.7 and 1,167 N501Y) and 4,299 were classified as non-VOC. The overall secondary attack rate was higher for VOC index cases (25.9%) compared to non-VOC (20.5%, p<0.01) with consistently higher secondary attack rates for VOCs across individual characteristics of the index cases (Table 1). The secondary attack rate of VOC index cases was 1.28 times higher than that of non-VOC index cases (RR=1.28, 95% CI 1.16-1.42); a similar trend of increased secondary attack rate was observed across subgroups (Figure 1).

**Table 1.** Characteristics of index cases by VOC status and household secondary attack rate

| Index case characteristics | Non-variant |  | Variant of Concern |  |
| --- | --- | --- | --- | --- |
|  | Index cases<br>No. (%) | Secondary attack<br>rate (%) | Index cases<br>No. (%) | Secondary<br>attack rate (%) |
| Total | 4,299 | 20.5 | 1,318 | 25.9 |
| Gender<sup>a</sup> |  |  |  |  |
| Female | 2,074 (48.2) | 19.8 | 647 (49.1) | 25.6 |
| Male | 2,214 (51.5) | 21.1 | 660 (50.1) | 26.2 |
| Age group |  |  |  |  |
| Median [IQR] | 39 [27, 55] |  | 37 [25, 52] |  |
| 0-9 years | 195 (4.5) | 17.0 | 55 (4.2) | 23.0 |
| 10-19 years | 351 (8.2) | 17.0 | 114 (8.6) | 20.1 |
| 20-29 years | 783 (18.2) | 15.8 | 319 (24.2) | 21.8 |
| 30-39 years | 837 (19.5) | 20.8 | 225 (17.1) | 25.2 |
| 40-49 years | 681 (15.8) | 20.4 | 233 (17.7) | 26.3 |
| 50-59 years | 711 (16.5) | 25.9 | 186 (14.1) | 34.9 |
| 60-69 years | 457 (10.6) | 25.3 | 126 (9.6) | 32.5 |
| 70-79 years | 188 (4.4) | 22.1 | 45 (3.4) | 24.3 |
| ≥80 years | 96 (2.2) | 21.9 | 15 (1.1) | 47.8 |
| Outbreak Related |  |  |  |  |
| No | 3,558 (82.8) | 20.7 | 1,040 (78.9) | 24.0 |
| Yes | 741 (17.2) | 19.0 | 278 (21.1) | 33.1 |
| Testing delay<sup>b</sup> |  |  |  |  |
| No symptoms | 410 (9.7) | 6.0 | 116 (8.9) | 12.4 |
| <0 days | 159 (3.8) | 6.3 | 71 (5.5) | 16.0 |
| 0 days | 431 (10.2) | 13.9 | 137 (10.5) | 15.5 |
| 1 day | 769 (18.2) | 21.6 | 258 (19.8) | 28.7 |
| 2 days | 672 (15.9) | 21.9 | 207 (15.9) | 33.2 |
| 3 days | 498 (11.8) | 23.9 | 138 (10.6) | 24.5 |
| 4 days | 335 (7.9) | 23.6 | 86 (6.6) | 31.4 |
| ≥5 days | 960 (22.7) | 26.2 | 288 (22.1) | 30.1 |
| Neighbourhood<br>Characteristics |  |  |  |  |
| Proportion of visible<br>minorities<sup>c</sup> |  |  |  |  |
| 0-20% | 1,128 (26.8) | 19.7 | 229 (17.6) | 24.0 |
| 21-40% | 659 (15.7) | 22.0 | 209 (16.1) | 26.8 |
| 41-60% | 649 (15.4) | 22.4 | 224 (17.2) | 21.7 |
| 61-80% | 765 (18.2) | 19.7 | 310 (23.9) | 29.3 |
| ≥81% | 1,001 (23.8) | 20.3 | 327 (25.2) | 26.3 |
| Proportion of households<br>with crowding<sup>d</sup> |  |  |  |  |
| < 5% | 2,381 (56.7) | 21.5 | 640 (49.3) | 27.4 |
| ≥ 5% | 1,821 (43.3) | 19.5 | 659 (50.7) | 24.6 |

<sup>a</sup>This section does not include missing or other genders (22 index cases and 20 associated secondary cases and 73 household contacts).
<sup>b</sup>Testing delay: days between the index case symptom onset and specimen collection. Cases with no symptoms were defined as cases that were missing symptom onset date and did not have any COVID-19 symptoms flagged in CCM. Index cases with a testing delay of <0 days were those who were tested prior to the onset of their symptoms (i.e. presymptomatic). Those that did not report a symptom or did not report a symptom onset date and were not reported
as asymptomatic, or did not report a specimen collection date, were excluded from this section (82 index cases and associated 59 secondary cases and 235 household contacts).
<sup>c</sup>Percentage of visible minorities (non-white and non-Indigenous population) in an Aggregate Dissemination Area having population between 5,000 to 15,000 based on the 2016 Census of Population in Canada. Households missing this measure were excluded from this section (116 index cases and associated 48 secondary cases and 311 household contacts).
<sup>d</sup>Percentage of households in an ADA which have more than 1 person per room. Households missing this measure were excluded from this section (116 index cases and associated 48 secondary cases and 311 household contacts).

**Figure 1.**
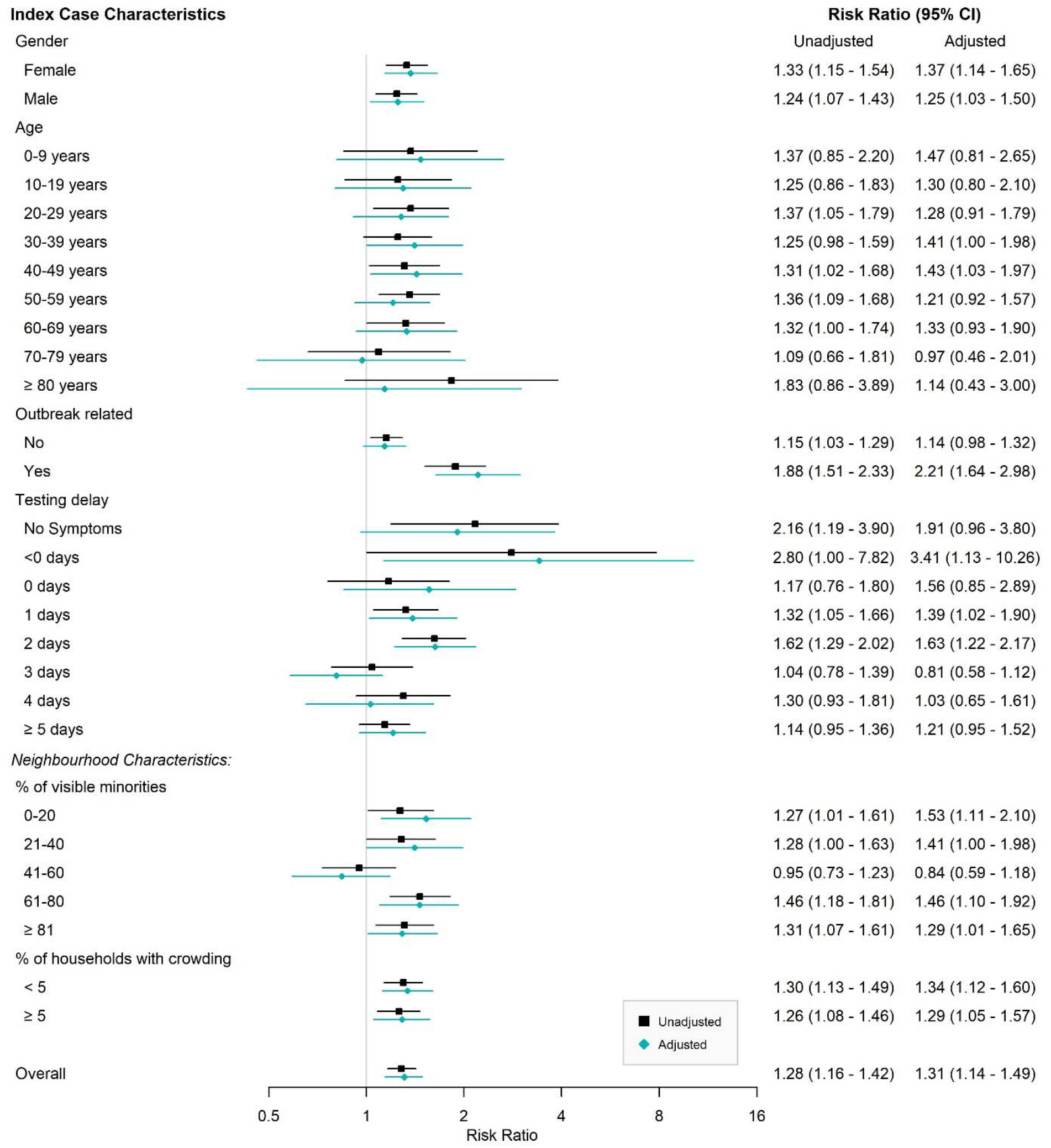
Risk ratio comparing household secondary attack rate associated with VOC vs. non-VOC index cases by characteristic of index case, unadjusted (black) and adjusted by 1:1 propensity-score matching (green) estimates Note: Unadjusted risk ratio accounts for household clustering and includes the full cohort. Adjusted risk ratio refers to the propensity score-matched cohort.

We included 1,259 index VOC and non-VOC cases in the propensity score-matched analysis. The secondary attack rate for VOC index cases in this matched cohort was 1.31 times higher than non-VOC index cases (RR=1.31, 95%CI 1.14-1.49), similar to the unadjusted estimate. In stratified analyses, the higher secondary attack rate for VOC compared to non-VOC index cases was accentuated for asymptomatic index cases (RR=1.91, 95% CI 0.96-3.80) and presymptomatic cases (RR=3.41, 95%CI 1.13-10.26)

## Discussion

In our cohort, we estimated that the household secondary attack rate of VOC index cases was 31% higher than non-VOC index cases, providing evidence of increased transmissibility. This is consistent with previous VOC (i.e., B.1.1.7) secondary attack rate estimates from the United Kingdom (relative secondary attack rate (32%, 12.9% vs 9.7% among all contacts);^6^ however, households are an important contributor to COVID-19 and provide a valuable setting in which to examine transmission.^7^ Our estimates of increased transmission from asymptomatic and pre-symptomatic VOC index cases has not previously been reported and suggests an increased importance of prevention measures when individuals are not aware of their infection. The biological mechanism responsible for increased transmissibility has not been identified, but hypotheses include higher viral loads (i.e. increased transmission potential per contact), and prolonged viral shedding (i.e., longer infectious period).^3^ The N501Y mutation has also been associated with enhanced binding affinity of severe acute respiratory syndrome coronavirus-2 (SARS-CoV-2) to angiotensin-converting enzyme 2 (ACE2) receptors.^13,14^

Limitations of this study include potential misclassification of secondary cases as index cases and small sample sizes in some subgroups. We may have underestimated secondary attack rates as we only captured diagnosed secondary cases and we lacked testing data on all household contacts; however, we do not believe this would be differential by VOC status of the index case. Ontario implemented more stringent measures for close contacts of all cases (not just VOC cases) in early February in response to VOC introductions, including increased frequency of testing during quarantine and outbreaks.^15^ If household contacts were more likely to test once VOC status was known, a similar increase in the risk ratio would be expected across symptom status, suggesting an independent effect of asymptomatic and presymptomatic VOC cases on transmission.

This study provides strong evidence of increased transmissibility in households due to VOCs and suggests that asymptomatic and pre-symptomatic transmission may be of particular importance for VOCs. Our study suggests that more aggressive public health measures will be needed to control VOCs. While measures effective for persons with unknown disease status such as physical distancing and masking may continue to be highly effective, measures focused on symptomatic individuals, such as public health contact tracing, may be increasingly ineffective unless extremely rapid. Ongoing research is needed to understand mechanisms of VOC transmissibility to curb their associated morbidity and mortality.

## Data Availability

Data sharing requests should be directed to Public Health Ontario.

